# Systematic Prioritisation of AI-Detected Chest X-ray Abnormalities for Optimised Triage of Suspected Lung Cancer Patients Lung Cancer Detection

**DOI:** 10.1101/2024.12.30.24319499

**Authors:** Rhidian Bramley, Anna Sharman, Rebecca Duerden, Sarah Lyon, Melissa Ryan, Elodie Weber, Louise Brown, Matthew Evison

## Abstract

**Objective:** This study aimed to establish a reproducible method for categorisation of the AI-detected chest X-ray (CXR) abnormalities that should be prioritised for urgent reporting to support faster lung cancer diagnosis. By selecting findings informed by cancer prevalence and clinical significance, we sought to maximise detection while maintaining a high negative predictive value (NPV).

**Materials and Methods:** Two cohorts of CXRs were evaluated: (1) a retrospective cohort of patients with confirmed lung cancer and abnormal CXRs, and (2) a prospective cohort of primary care referred CXRs from seven Greater Manchester trusts, with the AI system in shadow mode. The AI triage system (Annalise Enterprise CXR) evaluated the relative prevalence of 124 abnormalities, and prioritisation strategies were assessed using sensitivity, specificity, positive predictive value (PPV), and NPV.

**Results:** A total of 1,282 lung cancer patients were included in cohort 1. In cohort 2, the AI system processed 13,802 CXRs. Sensitivity was 95.87% (94.77%-96.97%) in cohort 1, and specificity was 79.11% (78.43%-79.79%) in cohort 2, with an NPV of 99.95%.

**Conclusion:** This study presents a systematic, reproducible method for prioritising AI-detected CXR abnormalities, balancing high sensitivity and NPV while minimising low-risk prioritisation. This approach provides a data-driven alternative to traditional methods relying solely on clinical judgement.

## Introduction

The rapid identification and prompt actioning of imaging, particularly chest x-rays (CXR) that suggest lung cancer, is crucial for improving patient outcomes and aligning with the National Optimal Lung Cancer Pathway[1]. For most patients, except those presenting with haemoptysis, the first diagnostic test for symptoms that could indicate lung cancer is a CXR [2]. These symptoms span a range of alternative diagnoses leading to an exceptionally high volume of CXR performed - over 8 million per year in England [3]. However, the incidence of lung cancer in CXR requested in primary care is just 1.4% [4]. This scenario creates substantial demand for CXR reporting in a context where radiology departments are stretched, with most CXR returning normal findings.

According to the 2023 Clinical Radiology Census Report, the NHS spent £276 million on outsourcing, insourcing and temporary locums to manage the overwhelming reporting demand, the highest expenditure on record [5]. The report also highlighted that this workforce shortages contribute to unacceptable diagnostic delays, including for patients undergoing CXR.

Artificial Intelligence (AI) has been proposed as one solution to these challenges. AI-driven tools can identify CXR findings suspicious for lung cancer and flag them for expedited reporting. Traditional approaches for AI-based prioritisation often rely heavily on clinical, focusing on a small subset of findings deemed significant. However, this method may be subjective, prone to bias and differences in clinical opinion, and may not always reflect the true prevalence of AI detected abnormalities across patient cohorts.

This study addresses the need for a more systematic, evidence-based approach to identifying which CXR findings should guide AI prioritisation. The primary objective is to establish a rapid and reproducible method for selecting AI-detected CXR abnormalities to triage studies with suspected lung cancer for urgent reporting, the overall goal of which is to enable faster diagnosis of lung cancer. Here, we present an approach developed in Greater Manchester that enhances selection through empirical data rather than intuition alone. This study provides robust evidence to guide prioritisation, addressing an important gap in real-world AI implementation.

A visual summary of the study design, methodology, and prioritisation strategies is provided in the accompanying infographic (see Supplementary Material).

## Methods

### Study Setting and Population

The study was conducted across seven acute care trusts in Greater Manchester. Two distinct patient cohorts were evaluated with inclusion criteria for each cohort outlined below:

1. **Cohort 1:** A retrospective cohort of 1,282 patients with confirmed lung cancer and an abnormal CXR between 1st June 2020 and 1st June 2022. These cases were curated for a parallel diagnostic accuracy study. Patients were included if they had a CXR within six months of their lung cancer diagnosis, with the CXR report identifying an abnormality that required follow-up.
2. **Cohort 2:** A prospective analysis was designed to include all primary care CXRs, performed between 1st June 2024 and 30th July 2024, with the AI system running in shadow mode. All included CXRs were GP-referred, collected consecutively, and represented the general population undergoing CXR.

This study was conducted as a service evaluation under the governance of the participating trusts and did not require formal ethical approval. All data were anonymised prior to analysis in accordance with NHS information governance standards and GDPR regulations. The project complied with the UK Policy Framework for Health and Social Care Research.

### Reference Standard

For cohort 1, every study was considered positive for lung cancer, owing to the definition of their inclusion criteria. For cohort 2, a 1% detectable lung cancer prevalence was assumed, in line with published studies and previous audits. The assumed prevalence alleviated the need for resource intensive ground truthing and accounted for the low prevalence finding in an otherwise normal cohort.

### AI System

The **Annalise Container CXR (version 2.1; Annalise-AI)** was deployed using the Sectra Amplifier Platform. This AI system can detect up to 124 distinct findings on CXR, offering a comprehensive assessment of abnormalities During prospective data collection, the AI system was deployed in shadow mode, meaning that the outputs were not visible to clinical staff and did not influence patient care or diagnostic decisions. This ensured that the evaluation posed no risks to patient safety while allowing for a robust assessment of the system’s performance. It analysed every CXR in both cohorts, generating an output for each of the 124 possible findings. The results were then anonymised and exported as comma-separated values (CSV) for further analysis.

Patients were not involved in the design or conduct of this study. As the study focused on evaluating the technical performance of AI in a shadow mode without direct patient impact, patient involvement was not deemed necessary.

### Selection of Findings

All findings deemed to be time-critical were included as a CRITICAL finding. For consideration of which findings to include to expedite lung cancer patients specifically, an Excel-based tool was developed to evaluate the prevalence of AI-detected abnormalities across the two cohorts. This tool has been made available as an open-source resource upon request, to support reproducible configuration in shadow mode at other institutions.

The ratio of findings prevalence in cohort 1 compared to cohort 2 was computed as a metric to facilitate finding selection. The AI findings were classed into 3 initial priority groups through consensus by the supplier and clinical leads.

1. CRITICAL – Findings that potentially require urgent action. These were to be prioritised independent of whether they related to cancer.
2. HIGH – Findings of clinical significance that may need to be prioritised.
3. STANDARD – Other findings considered not to warrant clinical prioritisation.

Three prioritisation strategies for cancer were developed to determine which of the initial high priority group should be prioritised to optimise the detection of cancer:

1. **Strategy 1**: The 19 AI findings with the highest prevalence ratio (>5), maximising specificity.
2. **Strategy 2**: All the HIGH findings with a prevalence ratio greater than 2, enhancing sensitivity.
3. **Strategy 3**: A final set of findings based on clinical judgement, balancing sensitivity and specificity.

A clinical reference group, including thoracic radiologists and chest physicians, reviewed the findings’ relative prevalence in both cohorts and refined the final set of findings for prioritisation.

### Statistical Analysis

The performance of the AI system was evaluated by calculating sensitivity, specificity, positive predictive value (PPV), negative predictive value (NPV), false positive rate (FPR), and false negative rate (FNR) for each strategy. Sensitivity was determined using cohort 1 (lung cancer patients), while specificity, PPV, and NPV were estimated using cohort 2 (general referral population). The Excel tool also calculated the number of CXRs needed to be reported to detect one cancer case to help set expectations for the AI’s clinical impact.

In cohort 1, all chest X-rays (CXRs) were from 1,282 patients with confirmed lung cancer visible on CXR, providing a clear standard to calculate sensitivity:

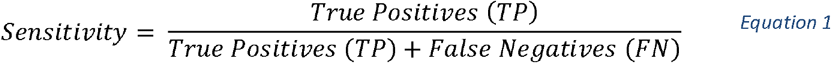

For cohort 2, a 1% detectable cancer prevalence was assumed based on published studies and previous audits of primary care CXRs, translating to an estimated 136 cancer positive cases in cohort 2 from a total 13,802 CXR. The True positives in this cohort were estimated as 136 x Sensitivity (computed from Cohort 1). The remaining cancer-negative cases (13,466) minus the false positives (FP) were used to calculate specificity:

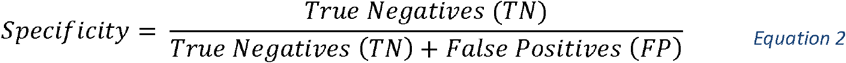

## Results

A total of 1,282 CXRs meeting the inclusion criteria were identified in cohort 1, and 13,802 CXRs meeting the inclusion criteria were included in cohort 2. Based on the assumed 1% lung cancer prevalence in cohort 2, this comprised 138 lung cancer positive cases and 13,664 lung cancer negative cases. The prevalence of all findings in each cohort and relative prevalences are shown in Supplementary Data (ST 1). The mean age in the cancer cohort was 71.4 years (SD ± 9.98), and the mean age in the referral cohort was 59.5 years (SD ± 17.4). There was no difference in the sex ratio between the cancer and referral cohorts, with 46.8% male and 53.2% female in each cohort. The flow of studies from each cohort is illustrated in Figure 1.

**Figure 1:**
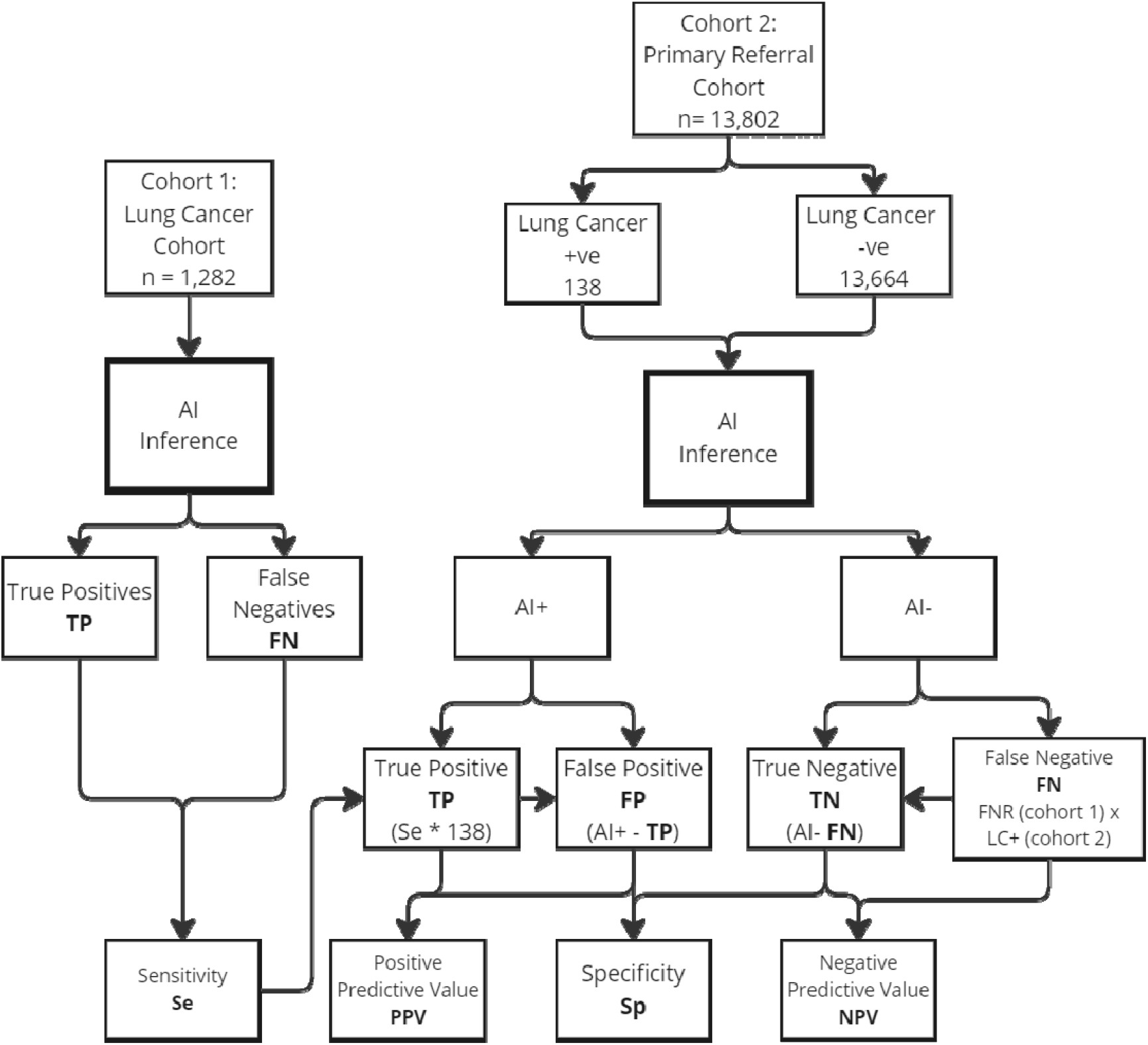
Flow diagram indicating inclusion of studies from each cohort, classification by the AI device and how the performance metrics were computed. The AI classifications varied based on the configuration of findings used.

The performance of the AI system across the three prioritisation strategies is summarised below and in Table 1: (see Supplementary Table 1 for full prevalence ratios):

**Table 1:** Performance metrics of the AI system across three prioritisation strategies for chest X-ray findings, including sensitivity, specificity, positive predictive value (PPV), and negative predictive value (NPV). PPV: Positive predictive value – the proportion of AI-positive cases that are true positives. NPV: Negative predictive value – the proportion of AI-negative cases that are true negatives. FPR: False positive rate – the proportion of non-cancer cases flagged by AI. FNR: False negative rate – the proportion of cancer cases missed by AI.

| Strategy | Strategy 1<br>(>5 ratio) | Strategy 2<br>(All HIGH) | Strategy 3<br>(Clinical Review) |
| --- | --- | --- | --- |
| AI Findings selected | 19 | 41 | 32 |
| Percent with AI finding<br>(Cohort 1) | 13.59% | 37.92% | 21.64% |
| Percent with AI finding<br>(Cohort 2) | 92.82% | 97.27% | 95.87% |
| True Positive (Cohort 1) | 1190 | 1247 | 1229 |
| False Negative (Cohort 1) | 92 | 35 | 53 |
| True Negative (Cohort 2) | 11916 | 8564 | 10809 |
| False Positive (Cohort 2) | 1748 | 5100 | 2855 |
| True Positive (Cohort 2) | 128 | 134 | 132 |
| False Negative (Cohort 2) | 10 | 4 | 6 |
| Sensitivity (95% CI) | 92.82%<br>(91.42% - 94.25%) | 97.27%<br>(96.37% - 98.18%) | 95.87%<br>(94.77% - 96.97%) |
| Specificity (95% CI) | 87.21%<br>(86.65% - 87.77%) | 62.68%<br>(61.87% - 63.49%) | 79.11%<br>(78.43% - 79.79%) |
| Positive predictive value (PPV) | 6.83% | 2.56% | 4.43% |
| Negative predictive value (NPV) | 99.92% | 99.96% | 99.95% |
| False Positive Rate (FPR) | 12.79% | 37.32% | 20.89% |
| False Negative Rate (FNR) | 7.18% | 2.73% | 4.13% |
| AI Positive Cancer Detection rate<br>(No CXR to report to detect 1 cancer) | 15 | 39 | 23 |
| AI Negative Cancer Detection rate<br>(No CXR to report to detect 1 cancer) | 1204 | 2274 | 1895 |

### Strategy 1

Prioritising the 19 AI findings classed as HIGH risk with a prevalence ratio greater than 5 for cancer resulted in a sensitivity of 92.82% and specificity of 87.21%. This strategy offered the highest specificity but at the expense of lower sensitivity.

### Strategy 2

Including AI findings classed as HIGH risk with improved sensitivity to 97.27% but reduced specificity to 62.68%.

### Strategy 3

The final set of findings, incorporating clinical judgement, resulted in a sensitivity of 95.87% in cohort 1, detecting 1,229 of the 1,282 patients with lung cancer. Specificity in cohort 2 was 79.11%, and the NPV was 99.95%.

To assess the impact of the assumed 1% cancer prevalence in Cohort 2,we conducted a statistical sensitivity analysis to determine how specificity varied at cancer prevalence rates ranging from 0.5%, to 2% in the referral cohort. Using prioritisation strategy 3, the resultant specificity ranged from 78.73% for a prevalence of 0.5% to 79.87% for a prevalence of 2%, which can be expressed as a confidence interval on the specificity based on the estimated cancer prevalence.

## Discussion

Prior to clinical use, the implementation of AI-driven triage devices relies on categorisation of the identifiable AI-findings to triage classes. The triage classes then dictate which studies are prioritised and the type of triage notification. The most common approach for this is either using the categorisation recommended by the vendor, or by clinical consultation at the deploying institution.

This study presents a novel and reproducible approach to prioritising CXR AI findings, combining empirical data with clinical judgement to optimise diagnostic accuracy. We employed a two-cohort design to assess both the sensitivity and specificity of the AI system for lung cancer detection on chest X-rays, addressing the challenges of using real-world data in a low-caner prevalence setting.

The cancer prevalence in patients referred from primary care according to NICE NG12 referral guideline in a large retrospective study was 1.4%[4]. The HealthCare Safety Investigation Branch has flagged that up to 20% of lung cancers are not detectable on CXR [6], so the number of detectable cancers is closer to 1% [7]

Calculating sensitivity required a retrospective cohort (cohort 1) with confirmed lung cancer cases, ensuring a sufficient number of verified positives to serve as the “ground truth”. This approach enabled a rigorous evaluation of the AI system’s ability to detect cancer in a cohort where all cases had a confirmed diagnosis visible on CXR, eliminating the need to wait for sufficient cancer cases to present from the referral population.

For specificity, we used a prospective cohort (cohort 2) in which the AI was run for a brief period in shadow mode on primary care CXRs. This approach allowed us to calculate specificity and understand the AI’s likely performance in identifying non-cancer cases within a referral population, thereby simulating the AI’s real-world impact without clinical risk.

By combining retrospective sensitivity with prospective prevalence estimates, this study design offered a safe and practical means to inform prioritisation strategies. This two-cohort method provides a valuable framework for assessing AI diagnostic tools prior to live clinical implementation, particularly in areas where the prevalence of the condition in the referral population is low.

The findings with the highest prevalence ratio in the HIGH priority list included solitary lung mass, inferior mediastinal mass, diffuse upper airspace opacity, diffuse airspace opacity, cavitating mass with content, hilar lymphadenopathy, and multiple masses or nodules. Interestingly, many bone related abnormalities (e.g. kyphosis, osteopaenia, spinal arthritis, and diffuse spinal osteophytes) also exhibited a higher prevalence in the lung cancer cohort. This was attributed to the difference in patient age between the cohorts, the average age in the cancer cohort more than 10 years over the referral cohort.

To explore this further, we conducted a supplementary age- and sex-matched analysis comparing the cancer cohort to a matched subset of the referral population. However, given that the matched cohort no longer represented the broader referral population and was not indicative of the number or proportion of CXRs that would be prioritised in practice, the results have not been included in the main findings. In current clinical deployment, AI triage tools do not support stratification by patient-level factors such as age, sex, or smoking history. Therefore, we focused on answering a practical clinical question: given a real-world referral population, which findings should be prioritised to maximise cancer detection while limiting over-prioritisation? The matched cohort analysis is available from the authors upon request.

Whilst the prevalence of findings in each cohort provides general insights, the overall objective was to identify a list of findings that would maximise the detection of suspected lung cancer cases, while ensuring critical findings were prioritised, and avoiding the unnecessary prioritisation of low-risk cases (thereby maintaining a high NPV). This study demonstrates a systematic approach to selecting AI-detected CXR findings for prioritisation in the context of lung cancer diagnosis. By analysing the prevalence of 124 findings in both a lung cancer cohort and a general referral population, we developed a data-driven method that maximises sensitivity while maintaining a high NPV.

**Strategy 1**, which focused only on the findings with the highest prevalence ratio, achieved high specificity but had a lower sensitivity. **Strategy 2**, which included all non-critical findings that may warrant prioritisation achieved the highest sensitivity but lowest specificity for cancer. **Strategy 3**, which balanced statistical prevalence with clinical judgement, offered the optimal combination of sensitivity and specificity, achieving a sensitivity of 95.87% and an NPV of 99.95%.

While this approach effectively prioritised cases with a high likelihood of cancer, the impact on non-prioritised cases warrants further exploration. Delays in reporting less critical findings could potentially affect patient outcomes for non-cancer conditions. Future studies should evaluate how prioritisation strategies impact overall diagnostic workflows and reporting times for all patients, including those with low-priority findings.

### Strengths

The approach proposed here differs from traditional methods by systematically prioritising findings based on their observed prevalence in cancer versus non-cancer cohorts, rather than relying solely on expert opinion. It is also reproducible across other ICBs implementing AI CXR prioritisation as part of the national AIDF programme. The methodology employed here can allow rapid completion of an initial study to support efficient implementation without the need for long term outcome data from prospective cohorts. The Excel workbook developed as part of this study was instrumental in refining our approach to AI finding prioritisation. By providing a platform to model different combinations of findings and instantly visualise their impact on diagnostic accuracy, this tool enabled a more data-driven and clinically relevant selection process. The tool is available on request and can be adapted by other sites to conduct a similar shadow mode analysis using their own local data. By inputting CSV files of AI findings for a cancer cohort and a referral cohort, users can explore how changes to the prioritisation configuration affect the proportion of patients flagged and cancers included. Future studies may benefit from adopting similar tools to streamline the integration of AI into clinical practice, particularly in settings where resource optimisation is critical.

### Limitations

The prioritisation system was developed and tested using data cohorts from patients within the same region, which introduces a risk of overestimating clinical performance without external validation. Additionally, the selected findings may not generalise to different geographical areas or populations.. However, a key strength of this approach is its adaptability, allowing the process to be replicated to identify relevant findings within local populations. Aggregated analyses of differences in prevalence could further enhance its applicability. External validation could be achieved by applying the tool to population cohorts outside Greater Manchester, which presents an opportunity for collaboration and further evaluation.

It is important to note that the performance metrics reported in this study are indicative rather than definitive. The reference standard for cohort 2 relies on an assumed prevalence, and analyses were conducted at the count level rather than validated against individual study-level reference standards. Consequently, some cases flagged as positive by the device may not be true positives, and some false positives could potentially represent true positives. An age- and sex-matched subgroup analysis was also conducted but not included in the final results, as the matched cohort did not reflect the characteristics of the full referral population or the real-world deployment context of the AI system.

However, the primary aim of this study was not to determine the diagnostic accuracy of the device but to identify findings that optimise sensitivity and negative predictive value (NPV). Expanding the inclusion of clinically relevant findings and applying the methodology to additional referral populations is expected to influence positive and negative predictive values. Nonetheless, the main objective was to select findings that maximise sensitivity and NPV for identifying suspected lung cancer cases.

Future directions include validating the findings and prioritisation strategies in diverse patient populations and healthcare settings. A priority is to conduct multicentre prospective clinical implementation studies to evaluate real-world performance, including operational impacts on reporting workflows. Collaborations with imaging networks are planned to ensure broader generalisability and external validation of the methodology.

Although patients were not directly involved in the design of this study, future work will incorporate patient perspectives, particularly regarding the ethical implications of prioritisation strategies. Engaging patients in the development of communication strategies for AI prioritisation will enhance transparency and trust in these technologies.

## Conclusion

This study presents a reproducible method for prioritising AI-detected CXR abnormalities, balancing the need for high sensitivity and NPV while accepting a lower PPV to avoid missing cancer cases and ensure early detection. By prioritising findings that maximise sensitivity, this approach improves the likelihood of identifying patients with suspected lung cancer. The use of shadow mode ensures clinical safety prior to full deployment, offering a practical, data-driven alternative to traditional judgement-based methods. With further validation, this approach could serve as a blueprint for integrating AI into radiology workflows, offering scalable solutions to global diagnostic challenges.

## Data Availability

All data produced in the present study are available upon reasonable request to the authors

## 1. Acknowledgements

The authors wish to thank the Greater Manchester Cancer Alliance, the participating NHS Trusts, and Annalise.ai for their support in this study. We also acknowledge Sectra Imaging IT Solutions for providing the platform for deploying the AI system.

## 9. Methodology

- Retrospective and observational study for cohort 1.
- Prospective and observational study for cohort 2.
- Multicentre study conducted across seven NHS Trusts in Greater Manchester.

## Key Points

### Key Point 1 - Question

#### Explain the unmet need/clinical problem your study addresses (20-25 words)

What is the best way to systematically prioritise AI-detected chest X-ray abnormalities to support faster and more accurate lung cancer diagnosis?

### Key Point 2 - Findings

#### Objectively summarize your main result (20-25 words)

This study demonstrated that prioritisation strategies can maximise sensitivity (95.87%) and negative predictive value (99.95%), while balancing specificity (79.11%) to ensure efficient lung cancer detection.

### Key Point 3 - Clinically Relevant Statement

#### Summarize the benefit for the patient and/or clinical relevance of the study (max. 40 words)

By prioritising clinically significant findings, this methodology improves early lung cancer detection, enhances reporting workflows, and supports scalable AI integration into radiology, benefiting patients and healthcare systems.

## Clinical Relevance Statement

The proposed prioritisation approach enhances early lung cancer detection by integrating AI into reporting workflows, addressing radiology demand and workforce constraints.

## Supplementary material ESM

**Supplementary Table 1 (ST1):**

| Initial Priority | AI Finding | Final priority | Count of studies (Cohort 1) | Count of studies (Cohort 2) | % of studies (cohort 1) | % of studies (cohort 2) | Prevalence Ratio (Cancer vs Referral) |
| --- | --- | --- | --- | --- | --- | --- | --- |
| Critical | Acute humerus fracture | P1 | 2 | 2 | 0.16% | 0.01% | 10.8 |
| Critical | Acute clavicle fracture | P1 | 1 | 1 | 0.08% | 0.01% | 10.8 |
| Critical | Widened aortic contour | P1 | 3 | 10 | 0.23% | 0.07% | 3.2 |
| Critical | Simple pneumothorax | P1 | 6 | 34 | 0.47% | 0.25% | 1.9 |
| Critical | Acute rib fracture | P1 | 17 | 228 | 1.33% | 1.65% | 0.8 |
| Critical | Pneumomediastinum | P1 | 0 | 1 | 0.00% | 0.01% | 0.0 |
| Critical | Tension pneumothorax | P1 | 0 | 4 | 0.00% | 0.03% | 0.0 |
| Critical | Subdiaphragmatic gas | P1 | 0 | 4 | 0.00% | 0.03% | 0.0 |
| Critical | Suboptimal ETT | P1 | 0 | 0 | 0.00% | 0.00% | 0.0 |
| Critical | Suboptimal NGT | P1 | 0 | 0 | 0.00% | 0.00% | 0.0 |
| Critical | Suboptimal CVC | P1 | 0 | 12 | 0.00% | 0.09% | 0.0 |
| Critical | Scapular fracture | P1 | 0 | 2 | 0.00% | 0.01% | 0.0 |
| Critical | Shoulder dislocation | P1 | 0 | 2 | 0.00% | 0.01% | 0.0 |
| Critical | Suboptimal PAC | P1 | 0 | 0 | 0.00% | 0.00% | 0.0 |
| Critical | Suboptimal gastric band | P1 | 0 | 2 | 0.00% | 0.01% | 0.0 |
| Critical | Subcutaneous emphysema | P1 | 1 | 0 | 0.08% | 0.00% | 0.0 |
| High | Solitary lung mass | P2 | 551 | 72 | 42.98% | 0.52% | 82.4 |
| High | Inferior mediastinal mass | P2 | 91 | 18 | 7.10% | 0.13% | 54.4 |
| High | Diffuse upper airspace opacity | P2 | 23 | 9 | 1.79% | 0.07% | 27.5 |
| High | Diffuse airspace opacity | P2 | 9 | 4 | 0.70% | 0.03% | 24.2 |
| High | Cavitating mass with content | P2 | 30 | 14 | 2.34% | 0.10% | 23.1 |
| High | Spinal wedge fracture | P2 | 18 | 9 | 1.40% | 0.07% | 21.5 |
| High | Hilar lymphadenopathy | P2 | 139 | 74 | 10.84% | 0.54% | 20.2 |
| High | Multiple masses or nodules | P2 | 160 | 87 | 12.48% | 0.63% | 19.8 |
| High | Perihilar airspace opacity | P2 | 13 | 8 | 1.01% | 0.06% | 17.5 |
| High | Lung collapse | P2 | 12 | 8 | 0.94% | 0.06% | 16.1 |
| High | Cavitating mass(es) | P2 | 64 | 53 | 4.99% | 0.38% | 13.0 |
| High | Diffuse lower airspace opacity | P2 | 54 | 55 | 4.21% | 0.40% | 10.6 |
| High | Solitary lung nodule | P2 | 411 | 474 | 32.06% | 3.43% | 9.3 |
| High | Segmental collapse | P2 | 181 | 210 | 14.12% | 1.52% | 9.3 |
| High | Loculated effusion | P2 | 31 | 39 | 2.42% | 0.28% | 8.6 |
| High | Focal airspace opacity | P2 | 639 | 825 | 49.84% | 5.98% | 8.3 |
| High | Multifocal airspace opacity | P2 | 104 | 144 | 8.11% | 1.04% | 7.8 |
| High | Spinal lesion | P2 | 1 | 2 | 0.08% | 0.01% | 5.4 |
| High | Pleural mass | P2 | 37 | 83 | 2.89% | 0.60% | 4.8 |
| High | Superior mediastinal mass | P2 | 56 | 131 | 4.37% | 0.95% | 4.6 |
| High | Diffuse pleural thickening | P2 | 15 | 37 | 1.17% | 0.27% | 4.4 |
| High | Upper interstitial thickening | P2 | 195 | 508 | 15.21% | 3.68% | 4.1 |
| High | Upper zone fibrotic volume loss | P2 | 53 | 157 | 4.13% | 1.14% | 3.6 |
| High | Rib lesion | P2 | 39 | 122 | 3.04% | 0.88% | 3.4 |
| High | Simple effusion | P2 | 289 | 953 | 22.54% | 6.90% | 3.3 |
| High | Diffuse nodular / miliary lesions | P2 | 20 | 68 | 1.56% | 0.49% | 3.2 |
| High | Lower zone fibrotic volume loss | P2 | 24 | 89 | 1.87% | 0.64% | 2.9 |
| High | Tracheal deviation | P2 | 75 | 285 | 5.85% | 2.06% | 2.8 |
| High | Diffuse interstitial thickening | P2 | 57 | 292 | 4.45% | 2.12% | 2.1 |
| High | Scapular lesion | P2 | 2 | 12 | 0.16% | 0.09% | 1.8 |
| High | Humeral lesion | P2 | 6 | 51 | 0.47% | 0.37% | 1.3 |
| High | Internal foreign body | P2 | 1 | 38 | 0.08% | 0.28% | 0.3 |
| High | Clavicle lesion | P2 | 0 | 9 | 0.00% | 0.07% | 0.0 |
| High | Calcified mass (> 5mm) | P3 | 23 | 126 | 1.79% | 0.91% | 2.0 |
| High | Pulmonary artery enlargement | P3 | 13 | 79 | 1.01% | 0.57% | 1.8 |
| High | Basal interstitial thickening | P3 | 169 | 1042 | 13.18% | 7.55% | 1.7 |
| High | Distended bowel | P3 | 5 | 50 | 0.39% | 0.36% | 1.1 |
| High | Widened cardiac silhouette | P3 | 170 | 2080 | 13.26% | 15.07% | 0.9 |
| High | Diffuse fibrotic volume loss | P3 | 2 | 41 | 0.16% | 0.30% | 0.5 |
| High | Pulmonary congestion | P3 | 16 | 333 | 1.25% | 2.41% | 0.5 |
| High | Peribronchial cuffing | P3 | 26 | 814 | 2.03% | 5.90% | 0.3 |
| Standard | Kyphosis | P3 | 23 | 6 | 1.79% | 0.04% | 41.3 |
| Standard | Osteopaenia | P3 | 21 | 8 | 1.64% | 0.06% | 28.3 |
| Standard | Spinal arthritis | P3 | 42 | 18 | 3.28% | 0.13% | 25.1 |
| Standard | Diffuse spinal osteophytes | P3 | 4 | 2 | 0.31% | 0.01% | 21.5 |
| Standard | Post resection volume loss | P3 | 19 | 60 | 1.48% | 0.43% | 3.4 |
| Standard | Lower zone bullae | P3 | 5 | 16 | 0.39% | 0.12% | 3.4 |
| Standard | Upper zone bullae | P3 | 27 | 88 | 2.11% | 0.64% | 3.3 |
| Standard | Rib resection | P3 | 3 | 12 | 0.23% | 0.09% | 2.7 |
| Standard | Reduced lung markings | P3 | 45 | 191 | 3.51% | 1.38% | 2.5 |
| Standard | Aortic stent | P3 | 2 | 10 | 0.16% | 0.07% | 2.2 |
| Standard | Diffuse bullae | P3 | 1 | 5 | 0.08% | 0.04% | 2.2 |
| Standard | Aortic arch calcification | P3 | 539 | 3016 | 42.04% | 21.85% | 1.9 |
| Standard | Gallstones | P3 | 3 | 17 | 0.23% | 0.12% | 1.9 |
| Standard | Calcified hilar lymphadenopathy | P3 | 3 | 17 | 0.23% | 0.12% | 1.9 |
| Standard | Hyperinflation | P3 | 318 | 1938 | 24.80% | 14.04% | 1.8 |
| Standard | Mastectomy | P3 | 25 | 158 | 1.95% | 1.14% | 1.7 |
| Standard | Coronary stent | P3 | 9 | 59 | 0.70% | 0.43% | 1.6 |
| Standard | Incompletely imaged chest | P3 | 108 | 717 | 8.42% | 5.19% | 1.6 |
| Standard | Calcified pleural plaques | P3 | 34 | 226 | 2.65% | 1.64% | 1.6 |
| Standard | Diaphragmatic elevation | P3 | 138 | 950 | 10.76% | 6.88% | 1.6 |
| Standard | Calcified axillary nodes | P3 | 5 | 35 | 0.39% | 0.25% | 1.5 |
| Standard | Shoulder replacement | P3 | 7 | 50 | 0.55% | 0.36% | 1.5 |
| Standard | Mediastinal clips | P3 | 42 | 305 | 3.28% | 2.21% | 1.5 |
| Standard | Axillary clips | P3 | 21 | 169 | 1.64% | 1.22% | 1.3 |
| Standard | Patient rotation | P3 | 305 | 2509 | 23.79% | 18.18% | 1.3 |
| Standard | Bronchiectasis | P3 | 25 | 209 | 1.95% | 1.51% | 1.3 |
| Standard | Hiatus hernia | P3 | 34 | 298 | 2.65% | 2.16% | 1.2 |
| Standard | Lung sutures | P3 | 4 | 37 | 0.31% | 0.27% | 1.2 |
| Standard | Atelectasis | P3 | 162 | 1529 | 12.64% | 11.08% | 1.1 |
| Standard | Electronic cardiac devices | P3 | 29 | 274 | 2.26% | 1.99% | 1.1 |
| Standard | Calcified granuloma (< 5mm) | P3 | 24 | 235 | 1.87% | 1.70% | 1.1 |
| Standard | Chronic rib fracture | P3 | 48 | 474 | 3.74% | 3.43% | 1.1 |
| Standard | Unfolded aorta | P3 | 321 | 3333 | 25.04% | 24.15% | 1.0 |
| Standard | Scoliosis | P3 | 74 | 810 | 5.77% | 5.87% | 1.0 |
| Standard | Chronic humerus fracture | P3 | 5 | 56 | 0.39% | 0.41% | 1.0 |
| Standard | Rotator cuff anchor | P3 | 7 | 81 | 0.55% | 0.59% | 0.9 |
| Standard | Sternotomy wires | P3 | 30 | 349 | 2.34% | 2.53% | 0.9 |
| Standard | Gastric band | P3 | 1 | 12 | 0.08% | 0.09% | 0.9 |
| Standard | Diaphragmatic eventration | P3 | 107 | 1310 | 8.35% | 9.49% | 0.9 |
| Standard | Breast implant | P3 | 5 | 62 | 0.39% | 0.45% | 0.9 |
| Standard | Nipple shadow | P3 | 24 | 301 | 1.87% | 2.18% | 0.9 |
| Standard | Chronic clavicle fracture | P3 | 8 | 102 | 0.62% | 0.74% | 0.8 |
| Standard | Shoulder fixation | P3 | 2 | 26 | 0.16% | 0.19% | 0.8 |
| Standard | Underexposed | P3 | 2 | 30 | 0.16% | 0.22% | 0.7 |
| Standard | Abdominal clips | P3 | 23 | 368 | 1.79% | 2.67% | 0.7 |
| Standard | Cervical flexion | P3 | 5 | 80 | 0.39% | 0.58% | 0.7 |
| Standard | Neck clips | P3 | 4 | 65 | 0.31% | 0.47% | 0.7 |
| Standard | Shoulder arthritis | P3 | 5 | 82 | 0.39% | 0.59% | 0.7 |
| Standard | Spinal fixation | P3 | 2 | 35 | 0.16% | 0.25% | 0.6 |
| Standard | Pericardial fat pad | P3 | 11 | 368 | 0.86% | 2.67% | 0.3 |
| Standard | Cardiac valve prosthesis | P3 | 2 | 88 | 0.16% | 0.64% | 0.2 |
| Standard | Underinflation | P3 | 1 | 90 | 0.08% | 0.65% | 0.1 |
| Standard | Airway stent | P3 | 0 | 1 | 0.00% | 0.01% | 0.0 |
| Standard | Clavicle fixation | P3 | 0 | 17 | 0.00% | 0.12% | 0.0 |
| Standard | Calcified neck nodes | P3 | 0 | 13 | 0.00% | 0.09% | 0.0 |
| Standard | Image obscured | P3 | 0 | 0 | 0.00% | 0.00% | 0.0 |
| Standard | Intercostal drain | P3 | 0 | 0 | 0.00% | 0.00% | 0.0 |
| Standard | In position CVC | P3 | 0 | 18 | 0.00% | 0.13% | 0.0 |
| Standard | In position NGT | P3 | 0 | 0 | 0.00% | 0.00% | 0.0 |
| Standard | In position ETT | P3 | 0 | 0 | 0.00% | 0.00% | 0.0 |
| Standard | Biliary stent | P3 | 0 | 3 | 0.00% | 0.02% | 0.0 |
| Standard | Oesophageal stent | P3 | 0 | 1 | 0.00% | 0.01% | 0.0 |
| Standard | In position PAC | P3 | 0 | 0 | 0.00% | 0.00% | 0.0 |
| Standard | Rib fixation | P3 | 0 | 2 | 0.00% | 0.01% | 0.0 |
| Standard | Pectus excavatum | P3 | 0 | 2 | 0.00% | 0.01% | 0.0 |
| Standard | Overexposed | P3 | 0 | 0 | 0.00% | 0.00% | 0.0 |
| Standard | Pectus carinatum | P3 | 0 | 0 | 0.00% | 0.00% | 0.0 |
| No Findings | NULL - no findings | P3 | 4 | 3350 | 0.31% | 24.27% | 0.0 |

### Matched Cohort Analysis

An age- and sex-matched analysis comparing the cancer cohort with a matched subset from the referral population was conducted but not included in the main results. These data are available from the authors upon request.

### Excel-based Prioritisation Tool

An open-source Excel workbook developed to support this analysis is also available from the authors upon request. The tool allows users to input CSV files of AI-detected findings from both a cancer cohort and a referral population to model different prioritisation strategies. It calculates how changes to the prioritisation configuration affect the proportion of cases flagged and the percentage of cancers included. The tool is designed to support reproducible local configuration of AI triage systems prior to clinical deployment in shadow mode.

### Infographic

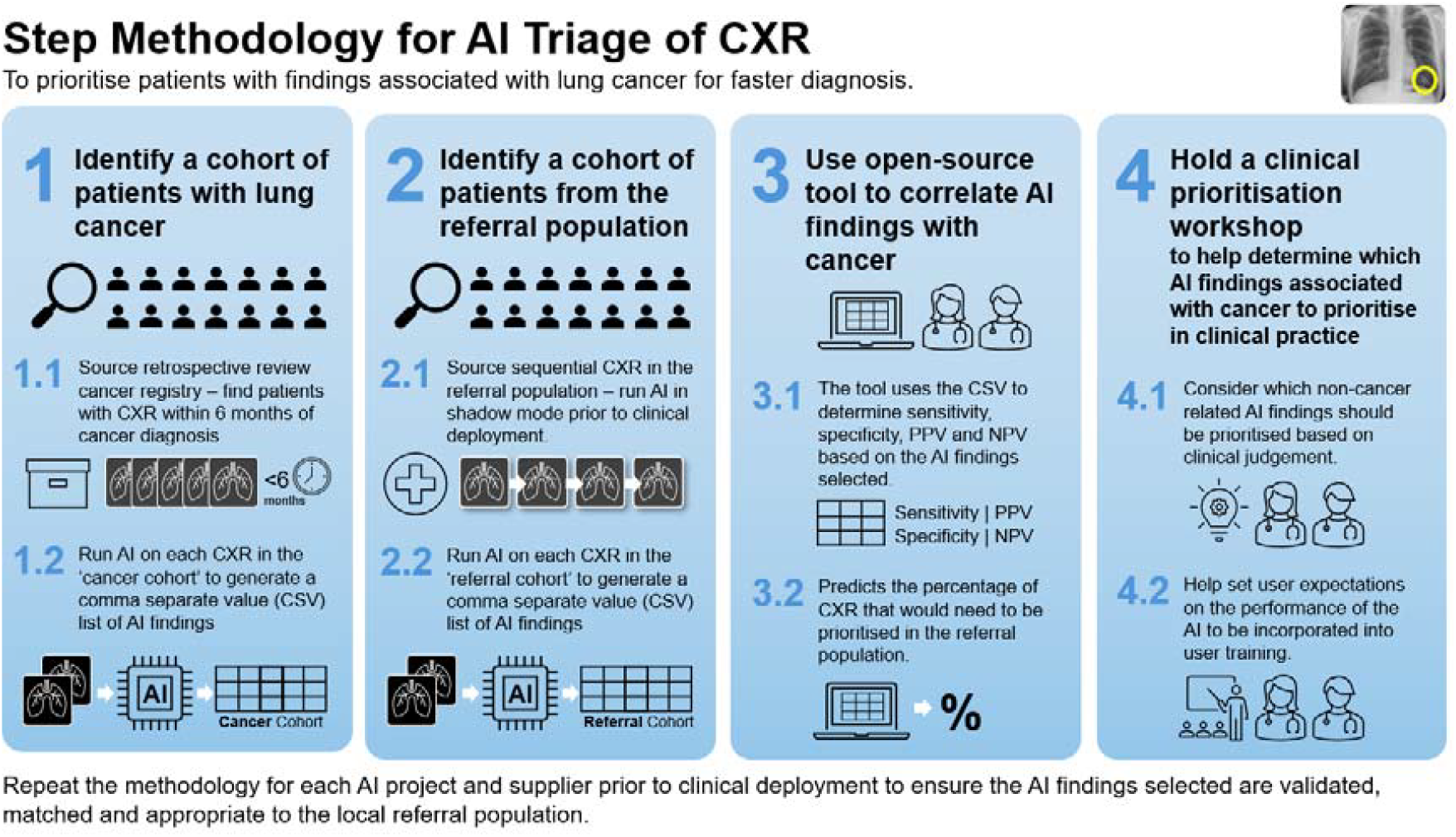

